# Does intimate partner violence associate with postnatal care utilization? Evidence from Tanzania’s 2022 Demographic and Health Survey

**DOI:** 10.64898/2026.09.01.26361973

**Authors:** Najma Ayoub Juma, Ramadhani M. Bofu, Joackim Kessy, Jean Burke

## Abstract

Postnatal care (PNC) is essential for reducing preventable maternal and neonatal deaths, but its utilization remain low across sub-Saharan Africa. Intimate Partner Violence (IPV) may be an under-recognized barrier to PNC utilization, particularly in Tanzania, where direct evidence shows that IPV is linked to limited utilization of PNC. Therefore, this study assessed the association between IPV and PNC utilization within 42 days postpartum among women in Tanzania. This study conducted a secondary analysis of the 2022 Tanzania Demographic and Health Survey (TDHS), a nationally representative cross-sectional survey. The analysis included 2,674 women aged 15-49 years who had a live birth in the five years preceding the survey and were selected for the domestic violence module. IPV (any, physical, sexual, and emotional) was the primary exposure, and PNC utilization within 42 days postpartum was the outcome. Modified Poisson regression was used to estimate crude and adjusted prevalence ratios (cPR/aPR) with 95% confidence intervals (CI) because the prevalence of the outcome was common. The prevalence of PNC utilization within 42 days postpartum was 42.0%, and the overall prevalence of IPV was 33.6% (physical 26.1%, emotional 21.8% and sexual 7.3%). Women who experienced any IPV had 16% lower PNC utilization than those who did not (aPR=0.84; 95% CI: 0.74-0.96). Physical IPV (16%, aPR=0.84; 95% CI: 0.73-0.96) and sexual IPV (25%, aPR=0.75; 95% CI: 0.57-0.98) were significantly associated with lower PNC utilization, while emotional IPV was not. Maternal education, partner’s age, travel time to the nearest health facility, and media exposure were also other covariates associated with PNC utilization. Intimate partner violence is associated with low utilization of PNC within 42 days postpartum in Tanzania. Integrating IPV screening and survivor support into postnatal care services, alongside addressing structural barriers to access, may improve postpartum care coverage and maternal-neonatal outcomes.

## Introduction

Maternal and newborn health remains a persistent global public health challenge. In 2023, a woman died from a pregnancy-related complication almost every two minutes worldwide, with over 90% of these deaths occurring in low- and lower-middle-income countries[1], and a substantial share occurring in the postnatal period[2]. Postnatal care (PNC) utilization is a critical strategy for reducing preventable maternal and neonatal deaths[3], yet many women do not receive recommended follow-up care[4]. Skilled postnatal checkups enable early detection and management of life-threatening complications such as postpartum haemorrhage, maternal infection, and neonatal sepsis[5,6].

In Tanzania, maternal mortality remains high at 104 per 100,000 live births and neonatal mortality at 24 per 1,000 live births[7], both exceeding Sustainable Development Goal targets 3.1 and 3.2. Despite investments in maternal health services, only 34% of women nationally receive PNC within two days of delivery[8], suggesting persistent barriers related to access, cost, distance, or quality of care.

Intimate partner violence (IPV) inflicted by a current or former partner[9] disproportionately affects women and may include controlling behaviors alongside physical, sexual, and emotional abuse[10]. IPV is known to reduce antenatal care attendance and is likely to affect postnatal care as well[11]; in Kenya, women who experienced IPV were significantly more likely to avoid postpartum healthcare altogether[12]. In Tanzania, IPV is highly prevalent during pregnancy and postpartum[13] and has emerged as a barrier to accessing PNC, contributing to reduced healthcare access and adverse maternal and newborn outcomes[14].

Globally, an estimated 27% of women have experienced physical or sexual IPV, or both, in their lifetime[15], with pooled lifetime prevalence estimates as high as 37.3%[16]. Regional estimates vary widely: 51.7% in a European sample[17], 6.1–67.4% across Asian settings[18], and 44% in sub-Saharan Africa[19]. In East Africa, pooled Demographic and Health Survey (DHS) analyses report IPV prevalence ranging from 32.7%[20] to 43.7%[21]. In Tanzania specifically, the 2015/16 TDHS reported 46% lifetime IPV prevalence among ever-married women[22], and localized studies in rural Tanzania report prevalence as high as 70.3% during pregnancy[23].

PNC coverage also remains inadequate globally, particularly in low- and middle-income countries. Reported PNC utilization ranges from 68.9% in Indonesia[24] to substantially lower levels across sub-Saharan Africa 27.4% pooled across 20 countries[25], 37% in Nigeria[26], 31.7% in East Africa[27], and 23.9% in rural southern Ethiopia[28]. In Tanzania, early PNC utilization rose from 37.4% in 2015/16[29] to 45% in 2022[30], indicating gradual improvement but continued need for targeted intervention.

Evidence on the association between IPV and PNC utilization is mixed and largely absent for Tanzania. A large multi-country analysis using DHS data from 36 low- and middle-income countries found physical IPV reduced antenatal and delivery care use but found no significant association with PNC utilization[31]. In contrast, studies in Pakistan[32], India[33], and pooled South Asian countries[34] found IPV was associated with reduced PNC and postnatal caregiving practices. Similarly, studies in Ethiopia[35,36] and Kenya[12] link IPV during the postpartum period to elevated maternal depression, anxiety, and stress that in turn impair health-seeking behavior, and Ethiopian national data show emotional IPV specifically linked to poor maternal healthcare utilization[37]. IPV has also been linked to poor maternal mental health, which may further reduce motivation to seek postnatal care[38].

Despite the high documented burden of both IPV and low PNC utilization in Tanzania, and the availability of nationally representative TDHS data with a domestic violence module, few studies have directly examined this association. This study addresses that gap by assessing the association between IPV and PNC utilization within 42 days postpartum among women in Tanzania, using the Andersen Behavioral Model[39] to frame IPV as a need factor influencing healthcare-seeking behavior alongside predisposing and enabling factors (Fig 1).

**Fig 1.**
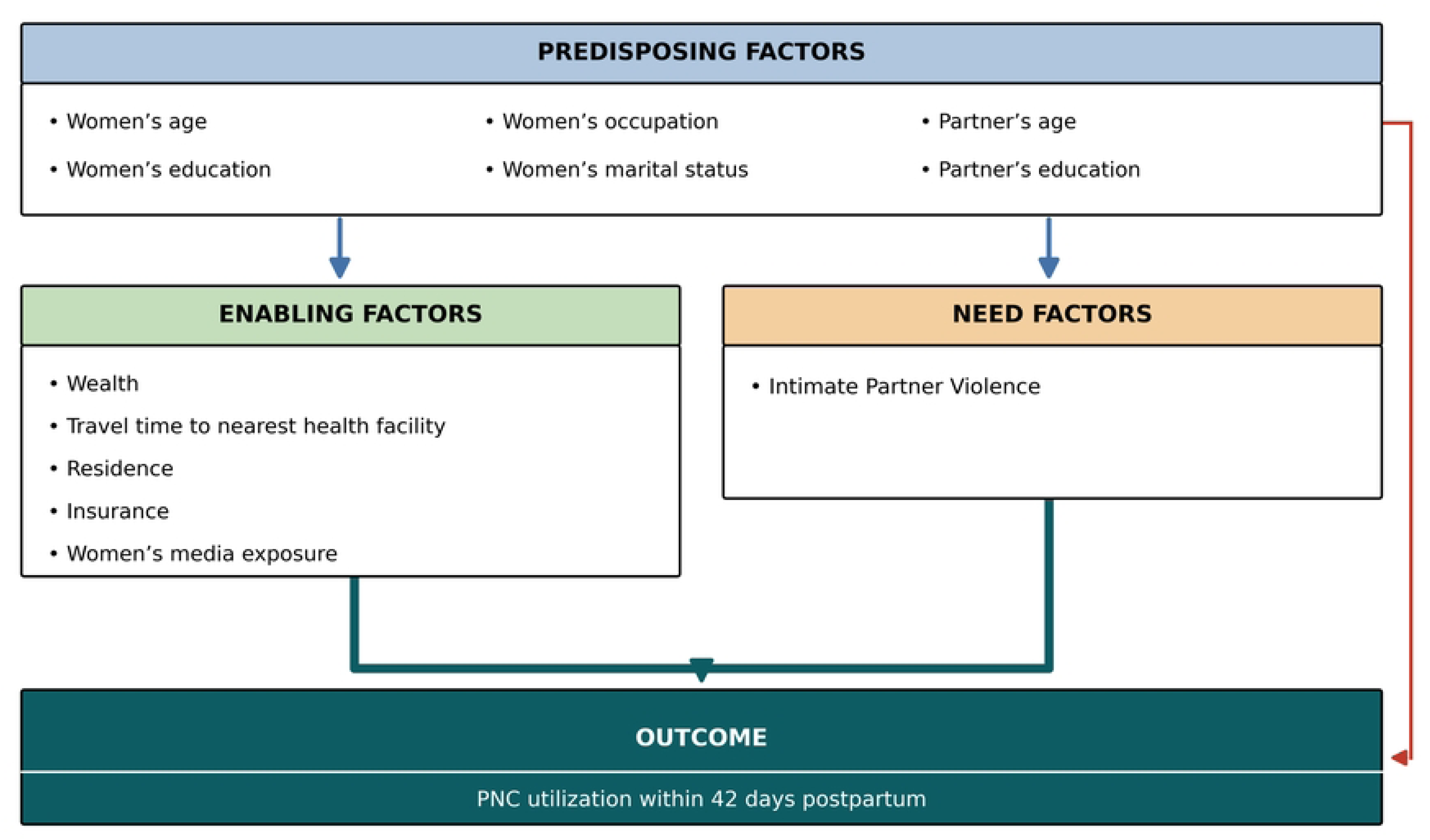
Modified conceptual framework of factors associated with postnatal care utilization.

## Methods

### Study design and data source

This study used data from the 2022 Tanzania Demographic and Health Survey (TDHS), a nationally representative cross-sectional survey conducted approximately every five years to provide reliable information on population and health indicators[30]. The TDHS employed a two-stage stratified cluster sampling design based on the 2022 Population and Housing Census. In the first stage, 629 clusters were selected (211 urbans, 418 rural); in the second stage, 26 households were systematically selected per cluster, yielding 16,354 households, of which 15,705 (99%) were successfully interviewed. Of 15,699 eligible women aged 15-49 years, 15,254 (97%) were interviewed[30]. Data were collected using standardized, pre-tested questionnaires adapted from the DHS Program’s Model Questionnaires, administered in Kiswahili through face-to-face interviews. The domestic violence module was administered privately to a randomly selected subsample of eligible women following WHO ethical guidelines.

### Study population and sample

This secondary analysis was restricted to women aged 15-49 years who had a live birth within the five years preceding the survey and who were selected for the domestic violence module. Women whose most recent pregnancy ended in stillbirth were excluded, as stillbirths are not eligible for PNC measurement in the TDHS. Of the 15,254 women interviewed in the 2022 TDHS, 5,563 responded to the domestic violence module; of these, 2,777 reported a live birth in the five years preceding the survey. After excluding 103 women with missing data on intimate partner violence, the final analytical sample comprised 2,573 women (weighted to 2,674 women to account for the complex survey design) (Fig 2). Using the method described by Kirkwood and Sterne[40], the study had 96.8% power to detect the observed effect size at a 5% significance level, based on a baseline PNC prevalence of 37.5% reported in prior literature[27].

**Fig 2.**
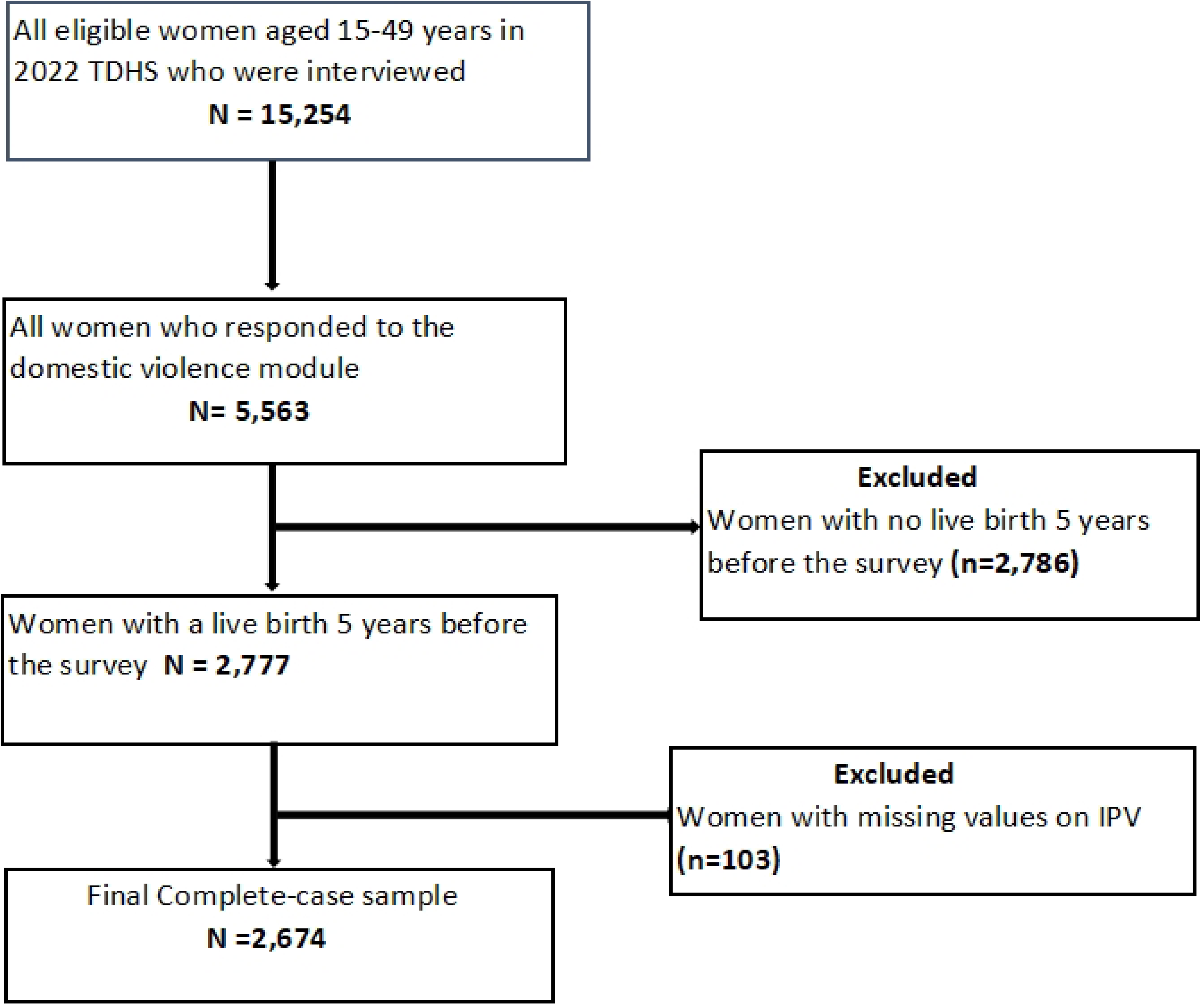
Flow chart for selection of study participants.

### Variables

The outcome variable was PNC utilization within 42 days postpartum, coded as a binary variable (received at least one postnatal check within 42 days = 1; none, or received after 42 days = 0), consistent with WHO and national guidelines[2]. The primary exposure was IPV, measured using the TDHS domestic violence module and coded as any IPV (yes/no), as well as three specific forms physical, sexual, and emotional analyzed separately and in combination. A woman was classified as having experienced any IPV if she reported at least one act of physical, sexual, or emotional violence by a partner. Covariates were selected based on the Andersen Behavioral Model[39] and prior literature, and included predisposing factors (women’s and partner’s age and education, marital status, occupation) and enabling factors (household wealth, travel time to the nearest health facility, residence, health insurance, media exposure).

### Data analysis

Analyses were conducted in Stata version 17, accounting for the TDHS’s multi-stage stratified cluster design using sampling weights, primary sampling units, and stratification. Descriptive statistics summarized baseline characteristics; weighted prevalence of IPV (overall and by type) and PNC utilization were estimated with 95% CIs. Chi-square tests assessed bivariate associations; variables with p<0.20 or documented clinical relevance were considered for multivariable analysis. A multilevel null model showed minimal between-cluster variance (intra cluster correlation coefficient = 0.016), which collapsed after adjustment for individual-level covariates, indicating no significant residual clustering. As log-binomial regression failed to converge, modified Poisson regression with robust (sandwich) variance estimation was used to estimate crude and adjusted prevalence ratios (cPR/aPR) with 95% CIs, given the binary outcome and its relatively high prevalence. Multicollinearity was assessed using variance inflation factors (all <10). Four adjusted models estimated associations for any IPV and each IPV form, each controlling for the covariates above. Potential interaction effects between IPV and key socio-demographic factors (education, wealth, and residence) were also examined to assess possible effect modification; no interaction terms were retained in the final models. Statistical significance was set at p<0.05.

Missing data were assessed during data cleaning; the proportion of missing values was below 5% and missingness was not associated with observed variables, so data were assumed to be missing completely at random. A complete-case analysis was therefore used, restricting the analytic sample to the 2,674 respondents with complete data on all key variables.

### Ethics statement

The study used secondary, de-identified data from the 2022 TDHS. Ethical clearance for the parent study and the domestic violence module was obtained by the DHS Program in accordance with WHO ethical and safety guidelines for research on violence against women; all original participants provided informed consent. Ethical clearance for this secondary analysis was obtained from the KCMC University Research and Ethics Committee, and permission to use the dataset was granted by the DHS Program.

## Results

### Socio-demographic characteristics of the study participants

A total of 2,674 women of reproductive age were included. About 72% of women lived in rural areas, 59% had primary education, and 72% were currently married. The ages of the participants range from 15-49 years, with a median age of 28 years and an interquartile range of 21-37 years. Of the 2,674 participants, 96% had no health insurance, and the median travel time to the nearest health facility was 30 minutes with an interquartile range 10-45 minutes (Table 1).

**Table 1.** Socio-demographic characteristics of the study participants (N=2,674)

| Variables | Frequency | Percent |
| --- | --- | --- |
| **Women's age (years)** |  |  |
| **Median (IQR)** | **28 (21-37)** |  |
| 15-24 | 754 | 28.2 |
| 25-34 | 1267 | 47.4 |
| 35-49 | 653 | 24.4 |
| **Women's education** |  |  |
| No education | 531 | 19.9 |
| Primary education | 1571 | 58.7 |
| Secondary and above | 572 | 21.4 |
| **Marital status** |  |  |
| Currently married | 1918 | 71.7 |
| Not currently married | 756 | 28.3 |
| **Women's working status** |  |  |
| Not working | 970 | 36.3 |
| Working | 1704 | 63.7 |
| **Partner's age (years)** |  |  |
| **Median (IQR)** | **30 (30-47)** |  |
| 15-29 | 694 | 26.0 |
| 30-44 | 1511 | 56.5 |
| 45+ | 469 | 17.5 |
| **Partner's education** |  |  |
| No education | 366 | 13.7 |
| Primary | 1699 | 63.5 |
| Secondary and above | 609 | 22.8 |
| **Household wealth index** |  |  |
| Lowest | 1075 | 40.2 |
| Middle | 512 | 19.1 |
| Highest | 1087 | 40.7 |
| **Residence** |  |  |
| Urban | 763 | 28.5 |
| Rural | 1911 | 71.5 |
| **Health insurance coverage** |  |  |
| No | 2562 | 95.8 |
| Yes | 112 | 4.2 |
| **Time to nearest HF** |  |  |
| **Median (IQR)** | **30 (10 - 45) minutes** |  |
| <30 minutes | 1161 | 43.4 |
| 30-59 minutes | 792 | 29.6 |
| 60-119 minutes | 445 | 16.7 |
| ≥2 hours | 276 | 10.3 |
| **Media exposure** |  |  |
| No | 970 | 36.3 |
| Yes | 1704 | 63.7 |

*IQR interquartile range.*

### Prevalence of Intimate Partner Violence

The overall prevalence of intimate partner violence among the 2,674 women of reproductive age was 33.6%. Specifically, physical IPV was the most common form reported by 26.1%, followed by emotional IPV at 21.8% and sexual IPV at 7.3% (Figure 3).

**Fig 3.**
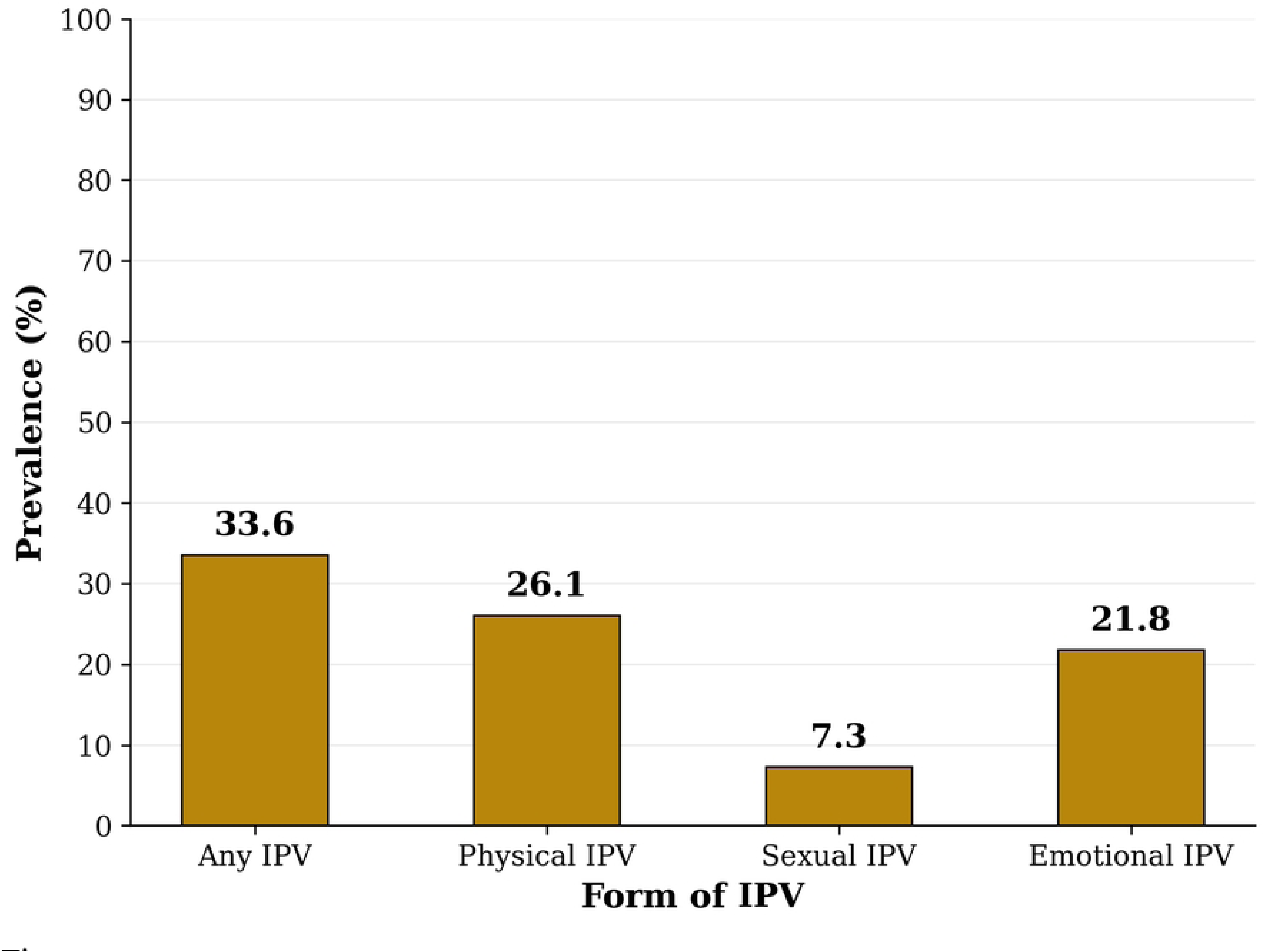
Prevalence of Intimate partner violence.

### Prevalence of Postnatal care utilization by IPV exposure and socio-demographic characteristics

Overall, 42.0% of the 2,674 women utilized the PNC within 42 days postpartum. The results showed significant differences in the prevalence of PNC utilization by any IPV, sexual IPV, physical IPV, women’s education, partner’s age and education, household wealth index, travel time to the nearest health facility, residence, media exposure and health insurance coverage.

The prevalence was 36.6% among women who experienced any form of IPV, 35.4% for physical IPV and 31.8% for sexual IPV. Also, the prevalence was 49.3% for women with secondary education or above, 50.6% for women whose partners had secondary education or above, 45.8% for women whose partners were aged 15-29 years, 50.2% for women from the highest wealth index, 7.4% for urban women, 59.8% for women with health insurance, 49.2% for those who took less than 30 minutes to the nearest health facility, and 46.8% for women with media exposure (Table 2).

**Table 2:** Prevalence of PNC utilization by IPV Exposure and Socio-demographic Characteristics (N=2,674)

| Variables | Total | PNC utilization (%) | P-value |
| --- | --- | --- | --- |
| **Any form of IPV** |  |  |  |
| No | 1775 | 794 (44.7) | **0.004** |
| Yes | 899 | 329 (36.6) |  |
| **Emotional IPV** |  |  |  |
| No | 2090 | 900 (43.0) | 0.116 |
| Yes | 584 | 223 (38.2) |  |
| **Sexual IPV** |  |  |  |
| No | 2480 | 1061 (42.8) | **0.038** |
| Yes | 194 | 62 (31.8) |  |
| **Physical IPV** |  |  |  |
| No | 1977 | 876 (44.3) | **0.001** |
| Yes | 697 | 247 (35.4) |  |
| **Women's age (years)** |  |  |  |
| 15-24 | 754 | 331 (43.9) | 0.316 |
| 25-34 | 1267 | 537 (42.4) |  |
| 35-49 | 653 | 255(39.1) |  |
| **Women's education** |  |  |  |
| No education | 531 | 166 (31.3) | **<0.001** |
| Primary | 1571 | 675 (43.0) |  |
| Secondary and above | 572 | 282 (49.3) |  |
| **Marital status** |  |  |  |
| Currently married | 1918 | 811 (42.3) | 0.750 |
| Not currently married | 756 | 312 (41.3) |  |
| **Women's working status** |  |  |  |
| Not working | 970 | 414 (42.7) | 0.734 |
| Working | 1704 | 709 (41.6) |  |
| **Partner's age group** |  |  |  |
| 15-29 | 694 | 318 (45.8) | **0.001** |
| 30-44 | 1511 | 652 (43.2) |  |
| 45+ | 469 | 153 (32.6) |  |
| **Partner's education** |  |  |  |
| No education | 366 | 125 (34.2) | **<0.001** |
| Primary | 1699 | 690 (40.6) |  |
| Secondary and above | 609 | 308 (50.6) |  |
| **Household wealth index** |  |  |  |
| Lowest | 1075 | 389 (36.2) | **<0.001** |
| Middle | 512 | 188 (36.7) |  |
| Highest | 1087 | 546(50.2) |  |
| **Residence** |  |  |  |
| Urban | 763 | 362 (47.4) | **0.005** |
| Rural | 1911 | 761 (39.8) |  |
| **Health insurance coverage** |  |  |  |
| No | 2562 | 1056 (41.2) | **0.001** |
| Yes | 112 | 67 (59.8) |  |
| **Travel time to nearest HF** |  |  |  |
| <30 minutes | 1161 | 571 (49.2) | **<0.001** |
| 30-59 minutes | 792 | 339 (42.8) |  |
| 60-119 minutes | 445 | 135 (30.3) |  |
| ≥2 hours | 276 | 78 (28.3) |  |
| **Media exposure** |  |  |  |
| No | 970 | 326 (33.6) | **<0.001** |
| Yes | 1704 | 797 (46.8) |  |

*P-values from Pearson's chi-squared test, Bold p-values are significant at $p < 0.05$*

### Association between IPV and PNC utilization within 42 days postpartum among women in Tanzania

The results from the modified Poisson regression adjusted analysis showed that, any IPV, sexual IPV and physical IPV were significantly associated with lower PNC utilization within 42 days postpartum, after adjusting for other variables. Women exposed to any IPV had 16% lower PNC utilization within 42 days postpartum than unexposed women (aPR=0.84; 95% CI: 0.74-0.96). Physical IPV was associated with a 16% lower PNC utilization (aPR=0.84; 95% CI: 0.73-0.96) and sexual IPV with a 25% lower PNC utilization (aPR=0.75; 95% CI: 0.57-0.98); emotional IPV showed no significant association. Other factors independently associated with PNC utilization within 42 days postpartum included primary education (aPR=1.21; 95% CI: 1.02-1.42, versus no education), media exposure (aPR=1.21; 95% CI: 1.06-1.38), partner’s age ≥45 years (aPR=0.74; 95% CI: 0.58-0.93, versus 15-29 years), and travel time to the nearest facility (60-119 minutes: aPR=0.68, 95% CI: 0.56-0.83; ≥2 hours: aPR=0.68, 95% CI: 0.53-0.87, versus <30 minutes) (Table 3).

**Table 3.**
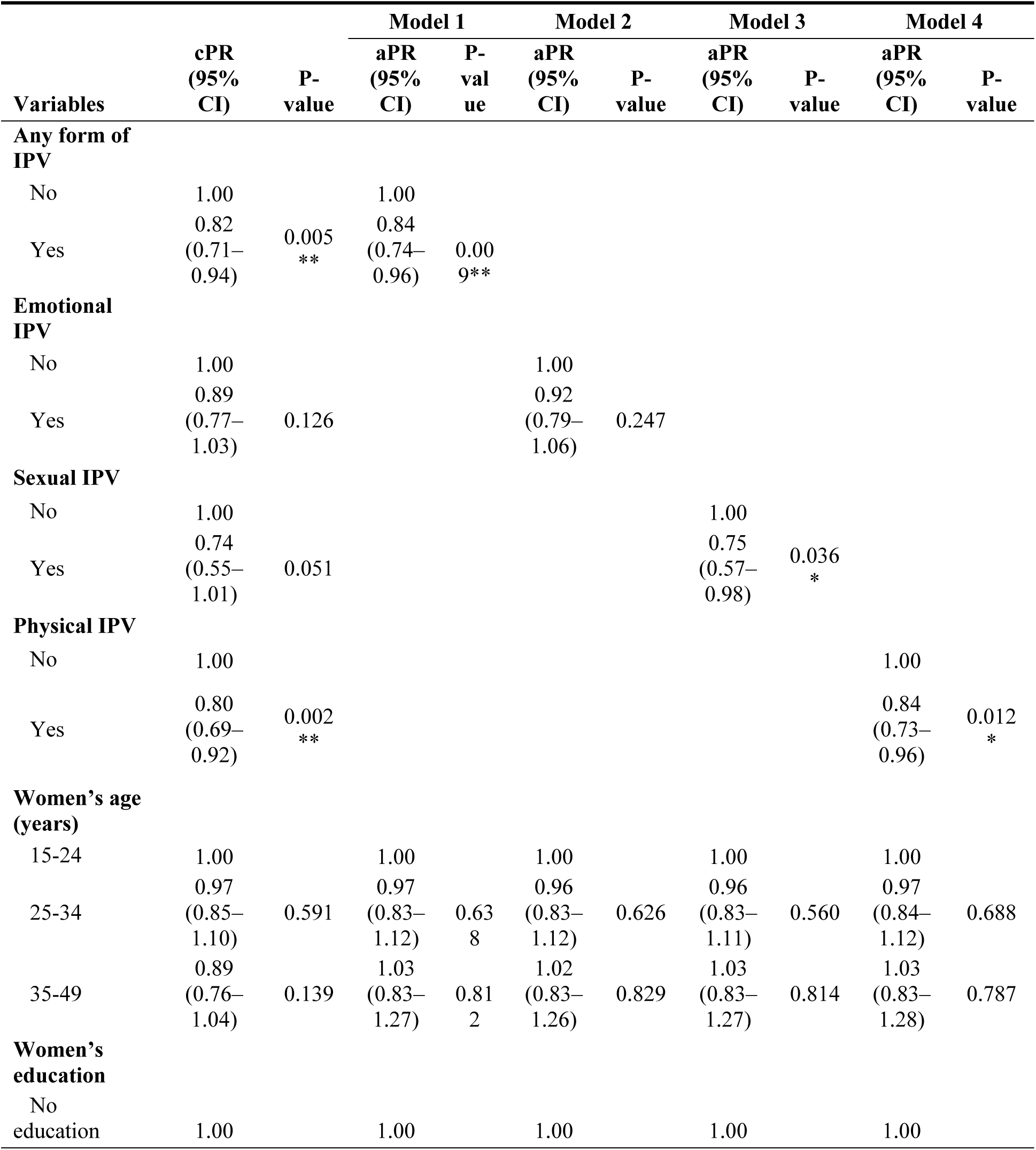
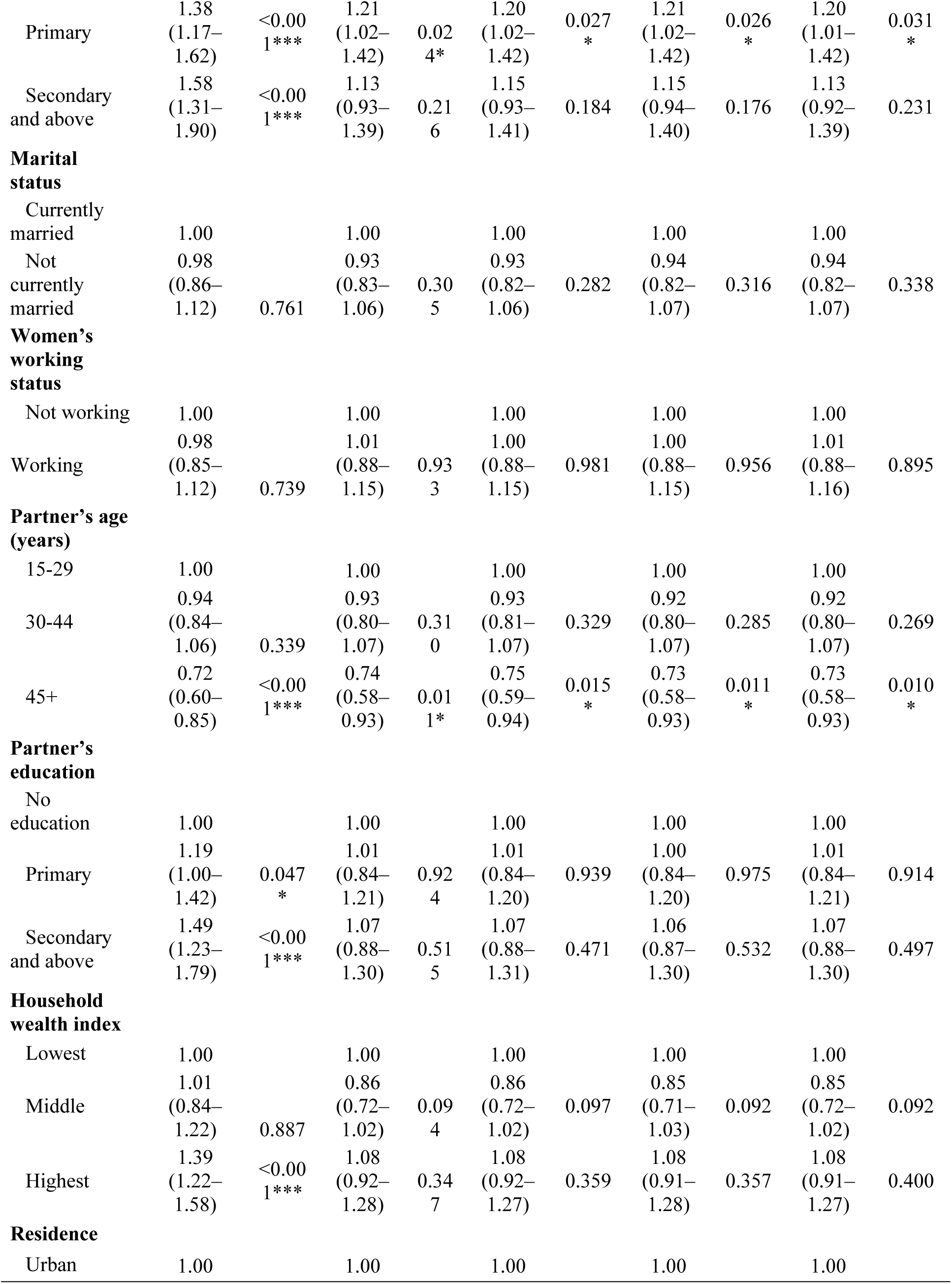
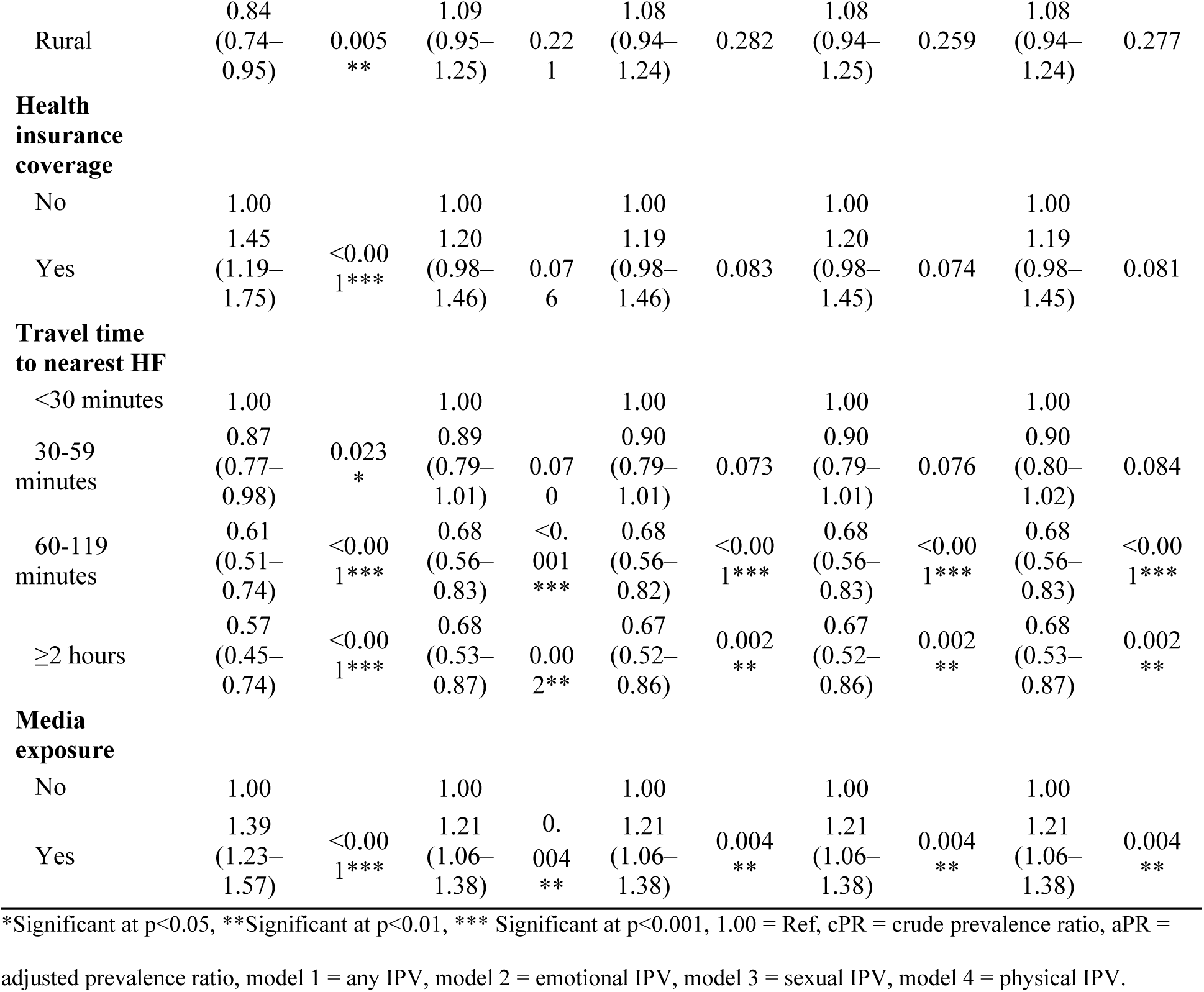
Association between IPV and PNC utilization among women in Tanzania (N=2,674)

## Discussion

This study found that 33.6% of women in Tanzania experienced IPV, and PNC utilization within 42 days postpartum was 42%. Women exposed to any IPV especially physical and sexual IPV were significantly associated with low PNC utilization within 42 days postpartum. Emotional IPV was not significantly associated with PNC utilization within 42 days postpartum.

The 33.6% prevalence of any IPV aligns with recent national estimates of 38.9%[7] and East African estimates of 32.7%[20], indicating a persistent threat to women’s health in Tanzania. Compared with the 2015/16 TDHS (46% overall)[22], this suggests a gradual decline, though levels remain high. Localized data from rural central Tanzania report substantially higher prevalence (70.3% during pregnancy)[23], reflecting geographic heterogeneity in patriarchal norms. The pattern of physical and emotional IPV being more common than sexual IPV mirrors regional findings[19,21], and the comparatively low reported sexual IPV likely reflects underreporting due to stigma and normalization of marital sexual coercion[23]. These findings support geographically targeted prevention, routine IPV screening, and culturally sensitive disclosure pathways integrated into maternal services, with prioritized outreach in high-burden rural areas[31,33].

PNC uptake within 42 days (42%) remained well below WHO recommendations[2]. This mirrors regional findings of inadequate PNC coverage, including 27.4% across 20 sub-Saharan African countries[25], with Eastern Africa at 31.7% and Tanzania near 37.5%[27], and low uptake among young women specifically (40%)[41]. Supply and demand-side barriers observed in Tanzania poor health literacy, overburdened staff, and supply shortages likely constrain uptake[42,29,43]. Strengthening facility capacity, improving supply chains, and expanding community-level education are needed to address both structural barriers and women’s information needs[27,42,29].

Consistent with the Andersen model’s need factors, IPV was associated with lower PNC utilization. Physical and sexual IPV likely impose immediate physical, psychological, and logistical constraints on care-seeking, while emotional IPV did not reach significance. This aligns with evidence that IPV undermines postpartum health practices[33] and reduces maternal healthcare access in Ethiopia and elsewhere[35,37,31]. Some studies report more complex associations[44], but multi-country analyses generally indicate IPV reduces service uptake, occasionally correlating with specific protective behaviors[45]. Similar patterns have been reported in Kenya and Pakistan[12,32]. The null finding for emotional IPV may reflect measurement limitations of standard survey instruments, which under-capture the severity and chronicity of psychological abuse[44]. As a modifiable barrier, IPV screening, referral pathways, and protective services should be incorporated into maternal health programs; health worker training to recognize and respond to IPV could improve retention across the postpartum continuum and reduce maternal-neonatal morbidity[12,45].

Beyond need factors, predisposing and enabling factors were also independently associated with PNC utilization, consistent with the Andersen model. Women with at least primary education were more likely to use PNC, consistent with multi-country evidence linking education to health literacy, decision-making autonomy, and awareness of postpartum monitoring benefits[27,41,44]. Women with older partners (≥45 years) were less likely to use PNC, possibly reflecting conservative gender norms limiting women’s autonomy; gender-transformative interventions engaging male partners may improve coverage[7,23]. Geographic access showed a graded effect, with women living 60–119 minutes or more than two hours from a facility having significantly lower PNC utilization, echoing regional findings[27,41,42]. Media exposure increased PNC use, underscoring the potential of targeted media campaigns; improving geographic access through decentralized services, outreach, and transport support, alongside mass and social media messaging, are recommended strategies[41,27].

### Strengths and limitations

This study used nationally representative TDHS 2022 data with rigorous multi-stage cluster sampling, supporting external validity, and applied modified Poisson regression with robust variance methodologically preferable to logistic regression for common binary outcomes while adjusting for complex survey design and key confounders. However, the cross-sectional design precludes causal inference; reliance on self-reported IPV and PNC data introduces potential recall and social desirability bias, which would most likely lead women to under-report violence and non-use of care, biasing the observed associations toward the null and making our estimates conservative; unmeasured confounders such as maternal mental health and facility-level quality of care were not captured; and the TDHS domestic violence module’s restriction to one randomly selected woman per household limited statistical power for rarer violence subtypes.

## Conclusions and recommendations

Physical and sexual violence, are associated with low PNC utilization within 42 days postpartum among women in Tanzania. Addressing IPV alongside structural barriers education, geographic access, and health communication is vital in improving postpartum care coverage and maternal-neonatal outcomes.

Policymakers need to expand PNC access in rural and hard-to-reach areas; the Ministry of Health need to consider integrating confidential IPV screening and referral pathways into postnatal guidelines so as to enable early identification of IPV survivors and timely linkage to protective and psychosocial support services, thereby improving PNC retention among women exposed to violence, healthcare providers should deliver trauma-informed, confidential care; and future researchers should pursue qualitative studies to understand how IPV shapes women’s autonomy and mobility in accessing postnatal care.

## Data Availability

No data was generated by this study. The following existing data sources were used: [The Tanzania Demographic and Health Surveys (TDHS) from [DHS Program] available via https://www.dhsprogram.com/data/dataset_admin/login_main.cfm

## Acknowledgments

The authors thank the DHS Program for granting access to the 2022 TDHS dataset.

